# Percutaneous Auricular Nerve Stimulation for Treating Post-COVID Fatigue (PAuSing-pCF)

**DOI:** 10.64898/2025.12.30.25343085

**Authors:** Maria Germann, Natalie J. Maffitt, Olivia A. Burton, Amn Ashhad, Anne M. E. Baker, Svetlana Cherlin, Marzieh Shahmandi, Norman Charlton, Aidan S. Baker, Boubker Zaaimi, Wan-Fai Ng, Demetris S. Soteropoulos, Stuart N. Baker, James M. S. Wason, Mark R. Baker

## Abstract

Even mild SARS-CoV-2 infection can lead to post-COVID syndrome, 70% of such patients have post-COVID fatigue (pCF). Many physiological abnormalities observed in pCF could be explained by reduced vagus nerve activity. The vagus nerve, central to metabolic and inflammatory homeostasis, can be activated non-invasively by transcutaneous auricular vagus nerve stimulation (taVNS). Can taVNS improve symptoms in pCF?

Data were collected from a randomized study including 114 individuals with pCF. They completed 16 weeks of daily home-based active, sham, or placebo taVNS. Data on subjective fatigue, captured by a Visual Analogue Scale (VAS), and objective measures of cortical excitability, muscle fatigue and autonomic function were collected.

In participants meeting minimum adherence (≥1 h/day on ≥50% of days), VAS and peripheral fatigue improved significantly after 8 weeks of active (but not sham or placebo) taVNS (11.9 ± 17.8 points improvement, p=0.003, N=24).

These results support taVNS as a potential therapy for pCF.

## Introduction

Long COVID has emerged as an unfortunate and common complication of the SARS-CoV-2/Corona Virus Disease 2019 (COVID-19) pandemic, affecting both those with severe disease requiring admission to hospital and those with relatively mild acute symptoms of COVID-19 infection. Currently, an estimated 3.1% of the UK population are experiencing long COVID symptoms^1^.

Studies show that at least 10–20% of all COVID-19-infected persons still suffer from long-term consequences of the disease six months after the initial infection^2^. These long-term sequelae are also referred to as post-COVID syndrome (PCS), which is defined by the National Institute for Care and Excellence (NICE) as symptoms that persist three months after the acute infection^3^.

Some individuals affected by PCS will spontaneously experience a relief of symptoms, or even a complete remission, after some time has passed since the onset of the disease^4^. However, many people continue to suffer from PCS, without any clear treatment or full understanding of the mechanisms behind the various long-term sequelae. Thus, long-term multi-organ consequences of SARS-CoV-2 infection remain an important clinical and scientific issue. Of the many symptoms of Long COVID, the commonest is fatigue; post-COVID fatigue (pCF) affects 71% of those with Long COVID in the UK^1^ which amounts to 2.3% of the UK population who suffered an acute COVID-19 infection.

Fatigue is a complex symptom, encompassing not only the perception of increased physical effort and extreme tiredness but also cognitive/mental fatigue (‘brain fog’). Muscle pathology has emerged as a common and debilitating feature, even in individuals with a history of mild COVID-19^5,6^. Reports highlight muscle weakness, myalgia, and impaired motor coordination among long COVID patients^7^, which can be disabling, and has a significant impact on quality of life.

These symptoms must emerge through changes at multiple levels of the peripheral and central nervous system. We previously hypothesized several plausible mechanisms by which immune or neuromodulator dysfunction could lead to changes in the nervous system and cause fatigue^8^. These relate to three broad areas: dysautonomia, cortical excitability and muscle pathology.

Whilst the severe acute respiratory syndrome coronavirus (SARS-CoV-2) which causes COVID-19 is not primarily neurotropic, neurological complications mediated by secondary coagulopathies or immune-mediated phenomena are common^9,10^. Such neurological complications include isolated or multiple cranial neuropathies; of these, vagus neuropathy is also recognized^11^ and may be a predictor of severe COVID-19^12^. The vagus nerve modulates immune processes via the cholinergic anti-inflammatory reflex^13^ and stimulation of the vagus nerve can activate central adrenergic/serotonergic^14^ and cholinergic^15^ pathways. A plausible hypothesis is that pCF is mediated, at least in part, by vagus nerve dysfunction.

Vagus nerve stimulation (VNS) is a well-established therapy (e.g. for intractable epilepsy), requiring surgical implantation of electrodes in the neck and a stimulator subcutaneously in the chest. More recently, novel approaches to non-invasive VNS stimulation (nVNS) have been developed. These use a handheld unit placed on the neck or stimulate the auricular branch of the vagus at the tragus of the external ear. The latter approach, transcutaneous auricular VNS (taVNS) has the advantage that it can be delivered using existing transcutaneous electrical nerve stimulation (TENS) devices. TENS is an established, affordable technology, available over the counter (or online) without prescription for self-administration without medical supervision.

Evidence that this approach might work to alleviate pCF comes from studies in patients with fatigue associated with primary Sjögren’s syndrome (a chronic autoimmune disorder), which showed rapid reductions in fatigue after 4-5 weeks of daily vagus nerve stimulation^16^. Similar positive findings were also reported in patients with fatigue associated with another chronic autoimmune disorder, systemic lupus erythematosus^17^. More recently, there have been studies using taVNS or cervical nVNS in patients hospitalized with acute COVID-19^18,19^ and small pilot studies have had some success in alleviating long-COVID symptoms^20,21^. However, to date larger-scale randomized and placebo-controlled studies evaluating the effects of patient self-administered taVNS on PCS have been conspicuous by their absence.

In this study we investigated the effects of taVNS, self-administered using a TENS device over 16 weeks, on symptoms and physiological, neurophysiological and behavioural correlates of fatigue in otherwise healthy people with pCF.

## Methods

### Participants

The study was approved by the Ethics Committee of Newcastle University Faculty of Medical Sciences. Participants provided written informed consent. The study was performed in accordance with the guidelines established in the Declaration of Helsinki. The trial was registered with the ISRCTN registry (ISRCTN18015802).

In total, 114 participants (85 females), aged 18-65, suffering from pCF by self-report were recruited. Only people scoring >=3 on a 0-10 Likert scale asking how fatigue affects their day-to-day living and answering yes to feeling that they needed treatment for their fatigue were invited to take part.

The diagnosis of COVID-19 was verified (by polymerase chain reaction, lateral flow or antigen tests) in all participants; none required hospitalisation following initial infection. Participants needed to have had ongoing symptoms of fatigue for at least 4 weeks post-infection, though most were several months or even years post-infection (see Figure 1).

**Figure 1:**
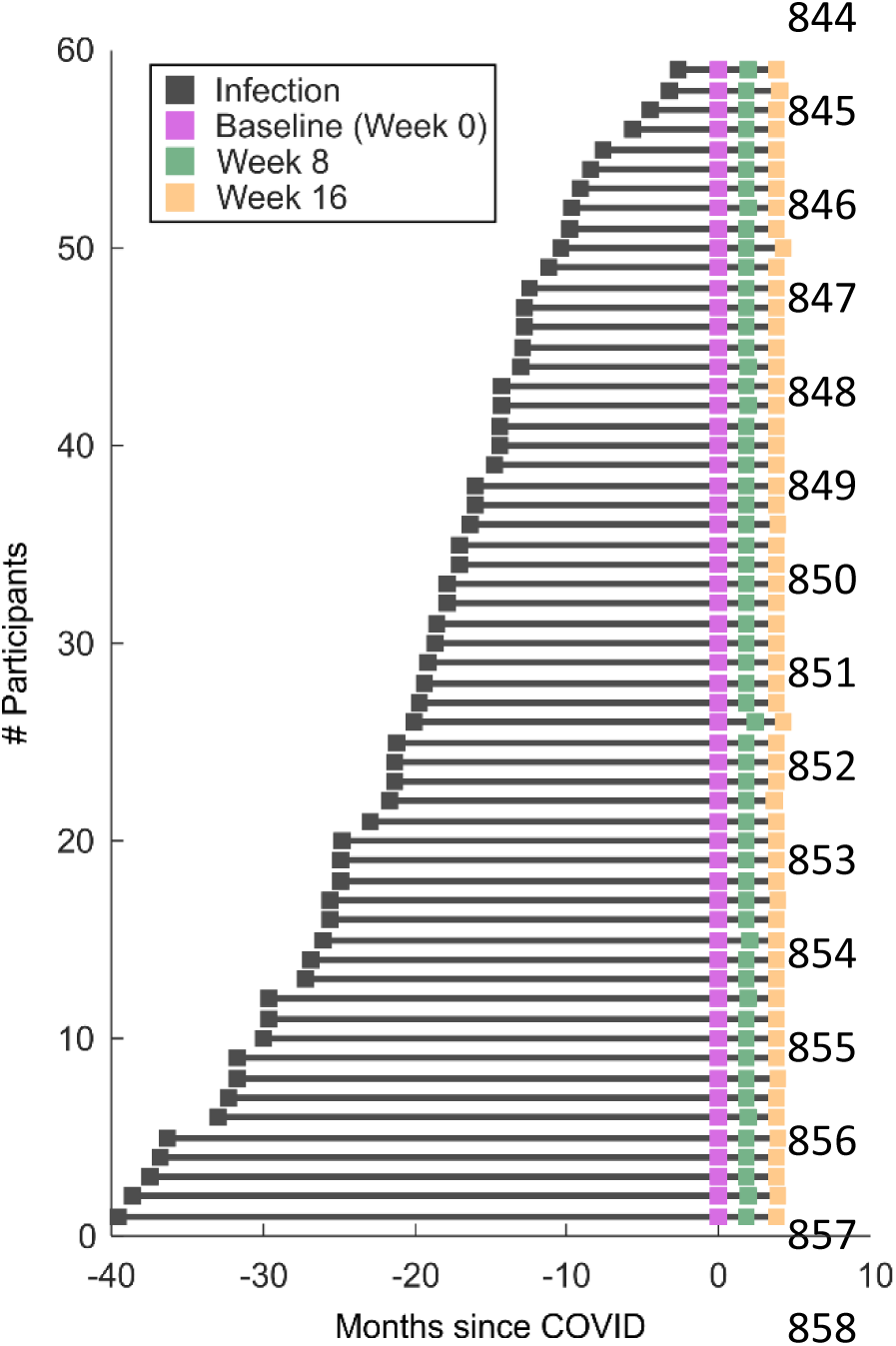
Distribution of times since the SARS-CoV-2 infection which caused persistent symptoms of fatigue in the pCF cohort.

After accounting for dropouts and participants not meeting minimum TENS usage (see Figure 2), data from 59 participants (45 females) were available for per protocol analysis and are presented here. None had a history of neurological or psychiatric disorders, nor cardiac disease prior to SARS-CoV-2 infection.

**Figure 2:**
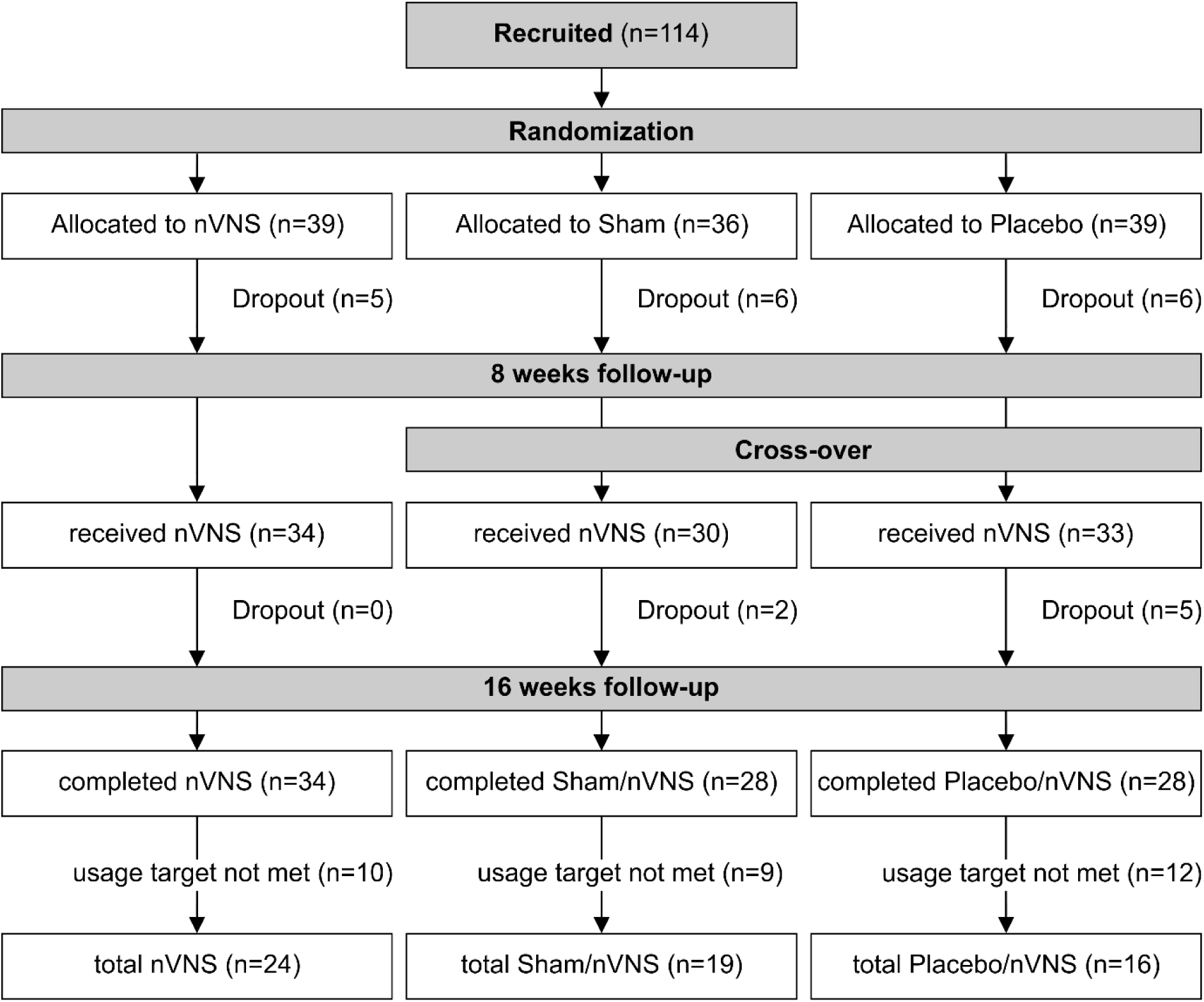
CONSORT diagram depicting participant flow through the study.

For the electrophysiological tests, the cohort of 52 healthy volunteers (37 females) with no symptoms of fatigue used in our previous papers^8,22^ served as the control dataset. In this control cohort, six subjects had reported having mild COVID-19 but with complete recovery and no symptoms of pCF.

### Study design

The study was a single-site, single-blind, sham-controlled and placebo-controlled, randomized trial design, with all participants moving to the active intervention (open label) after 8 weeks. The study design is summarised in the Consort diagram in Figure 2.

Once eligibility was confirmed, consent was taken, and baseline fatigue assessments and biometric, neurophysiological and behavioural assessments were completed (week 0). At baseline, participants were randomized to one of three experimental groups (see below) - active tragus stimulation, sham tragus stimulation (sham; control 1) and active pinna/greater auricular nerve stimulation (placebo; control 2). Participants were instructed not to use the stimulation for the first 7 days.

The main between-group results of the randomised study following the Statistical Analysis Plan are to be reported elsewhere. This paper reports a subgroup analysis which only included participants meeting minimum adherence (≥1 h/day on ≥50% of days), to explore causality and efficacy of the intervention.

Consumables and instructions for the assigned intervention were provided in a box. At week 8, participants returned to the lab and neurophysiological, behavioural and fatigue assessments were repeated, and participants were provided with a new box of equipment, consumables, and instructions for the final phase of the intervention. Participants receiving active VNS continued for a further 8 weeks and those initially in the control arms crossed over to the active intervention for the last 8 weeks of their participation in the study. This gave all participants the opportunity to benefit from the active taVNS intervention. At week 16, all assessments were repeated in a final visit.

Assessments were completed in the lab and one session took approximately 2 hours to complete.

### VNS stimulation

Participants were required to self-administer electrical stimulation to the external ear to activate nerves running beneath the surface of the skin. A FlexiStim (TensCare, Epsom, UK) transcutaneous electrical nerve stimulation (TENS) device was used for this purpose (Figure 3Aa). These devices are commercially available over the counter, or on-line, without prescription.

To activate the auricular branch of the vagus nerve (taVNS) a clip electrode (Figure 3Ac), manufactured in the lab and connected to the TENS system, was attached to the left tragus. Fabric sleeves, cut to the correct length to cover the two prongs of the ear clip electrode, ensured that the saline used to moisten the sleeves made a good electrical contact with skin overlying the tragus. To activate the greater auricular nerve (C2/C3 nerve roots; placebo control), the clip electrode was attached to the left earlobe/pinna (Figure 3Ad).

The stimulation parameters were set for each participant using the electrical muscle stimulation (EMS) program on the TENS machine and the same parameters were used for tragus and earlobe/pinna stimulation. The stimulation protocol began with a two-minute warm up at a frequency of 6Hz and pulse width of 200μs. This was followed by a 60-minute train, which alternated 30 seconds at 25Hz and 300μs with 60 seconds at 4Hz and 200μs. Finally, there was a 3-minute cool down stimulation protocol of 3Hz and 200μs. The stimulus current strength was set just above perceptual threshold to ensure the subject was aware of the stimulation, but also so that the intensity was not uncomfortable or painful.

To ensure participants were blinded, they were only required to power the TENS device ON (stimulator settings were pre-set for each participant and the same settings used for all interventions). A small plastic junction box (see Figure 3Aa) connecting the TENS cables to the earclip cables contained either a 10 MΩ resistor for active stimulation or a 10 Ω resistor to shunt current (Figure 3B). The resistor boxes were indistinguishable for participants. The experimenter was not blinded to the intervention after randomization.

**Figure 3:**
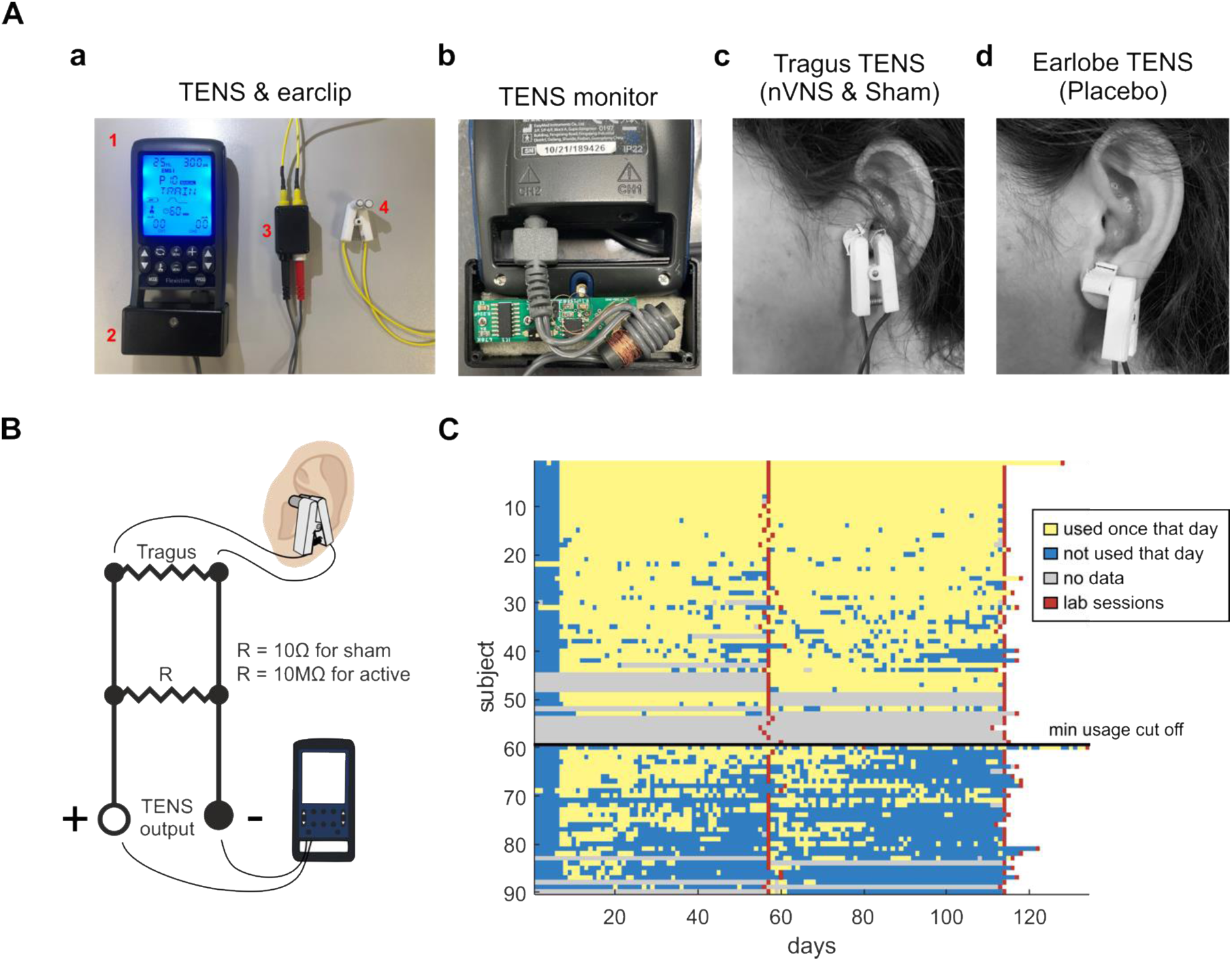
TENS equipment and adherence. **A:** TENS setup for taVNS. **(a)** Equipment consisted of *1*, a TENS machine, *2*, attached box to hold usage monitor, *3*, black box containing either active or sham resistor, *4*, earclip. **(b)** Open box from Aa showing usage monitor inside. **(c)** Earclip position on the tragus for active nVNS or sham stimulation. **(d)** Earclip position on the earlobe for placebo stimulation. **B:** Circuit diagram for resistor boxes (see 3Aa). For sham stimulation, a resistor of 10Ω was used to shunt the current. For active stimulation (active nVNS or placebo), a resistor of 10MΩ was used, which allowed the current to pass through the electrodes. **C:** TENS usage for each participant, with participants ordered in descending manner based on their usage. Each square represents usage for a day. Yellow indicates that taVNS has been used for at least 1h that day (corresponding to one TENS program cycle). Blue indicates that taVNS has not been used that day, or used for less than 1h that day. Grey indicates that there was no monitor data for those subjects due to technical issues. Red squares represent the day of the follow-up lab visits. Note that participants were instructed not to use the TENS during the first week, which is why all participants (with a few exceptions) show no usage for the first 7 days.

The three interventions were as follows:

#### Intervention-1 (active nVNS)

Clip electrode was attached to the tragus (Figure 3Ac) and connected to the TENS device to stimulate the auricular branch of the vagus nerve. The junction box connecting the TENS cables to the earclip cables contained a 10 MΩ resistor. Because this is much higher than the electrode resistance, almost all current flowed through the electrode, giving active stimulation (Figure 3B).

#### Intervention-2 (sham control)

Clip electrode was also attached to the tragus, but the junction box contained a 10 Ω resistor. Because this was much smaller than the electrode resistance, it shunted almost all of the stimulus current (Figure 3B). The auricular nerve was therefore not stimulated, but this condition controlled for mechanical pressure exerted by the tragus clip.

#### Intervention-3 (placebo control)

Clip electrode was attached to the earlobe (Figure 3Ad) and connected to the TENS device thus stimulating afferents of the greater auricular nerve (C2/C3 nerve roots). Junction box contained a 10 MΩ resistor for active stimulation (Figure 3B). This condition controlled for non-specific effects of electrical stimulation.

Before the participant started the allocated intervention, the experimenter also explained to each participant that, ‘because different current strengths are being tested, they may not feel the stimulation’.

Participants were asked to apply the intervention three times per day. Minimum usage was defined as using the TENS at least 1h a day (the length of one stimulation cycle) on at least 50% of days.

To monitor adherence, a data logger was attached in a plastic box to the TENS device (see Figure 3Ab). The battery-powered data logger comprised a microcontroller, electrically erasable programmable read-only memory and a ferrite rod around which the TENS cable was coiled. Every 10 minutes, the data logger powered up and determined if the TENS device was stimulating (determined by detecting the induced magnetic field from the current in the TENS cable). Every 4 hours the data logger wrote the count of ‘on’ states (between 0 and 24) to the memory. The data logger was powered by a coin cell battery, which allowed usage for the entire 8 week stimulation period without a battery change. At the end of the usage, count data could be downloaded to a personal computer and used to determine how often the TENS device had been used (Figure 3C). More technical details and a circuit diagram are provided in Supplementary File 1.

### Adverse Events

Overall, the stimulation was well tolerated, though many participants found the earclip to be mildly uncomfortable when it had to be worn on the tragus. No serious adverse events were reported during the study. Three people could not tolerate the pressure from the earclip when having to attach it to the tragus. Two people reported skin irritation at the area of stimulation, which could be treated with emollient cream and resolved after the study. Three people reported nerve pain in their neck, which also radiated to their arms or caused headaches. Only one of these participants was receiving active nVNS at the time, the other two were receiving placebo stimulation to the earlobe. Six participants reported a notable worsening of their fatigue over the course of the first 8 weeks of the study; these divided equally, with two participants receiving each type of stimulation. Out of the 12 people reporting adverse events, 5 decided to drop out of the study as a direct consequence, all of whom believed the stimulation made their fatigue worse and reported no other adverse events. Only one of these participants was receiving active nVNS at the time of drop out.

### Assessment procedures

#### Questionnaires

For patient-reported outcomes (PROs), participants were asked to fill out questionnaires online, over the course of the 16-week trial. Reminders were sent out as text messages containing the link to fill out the correct questionnaire on their mobile telephone.

The following short questionnaires were delivered every other day: Fatigue Visual Analogue Scale (VAS) and Functional Assessment of Chronic Illness Therapy Fatigue Scale (FACIT-F).

Once a week, participants were additionally asked to fill out a Fatigue Impact Scale (FIS).

#### General electrophysiological methods

The assessment protocols follow our previous study^8^ which compared participants with pCF to age-matched controls. A summary of the various electrophysiological measures is illustrated in Figure 4.

**Figure 4:**
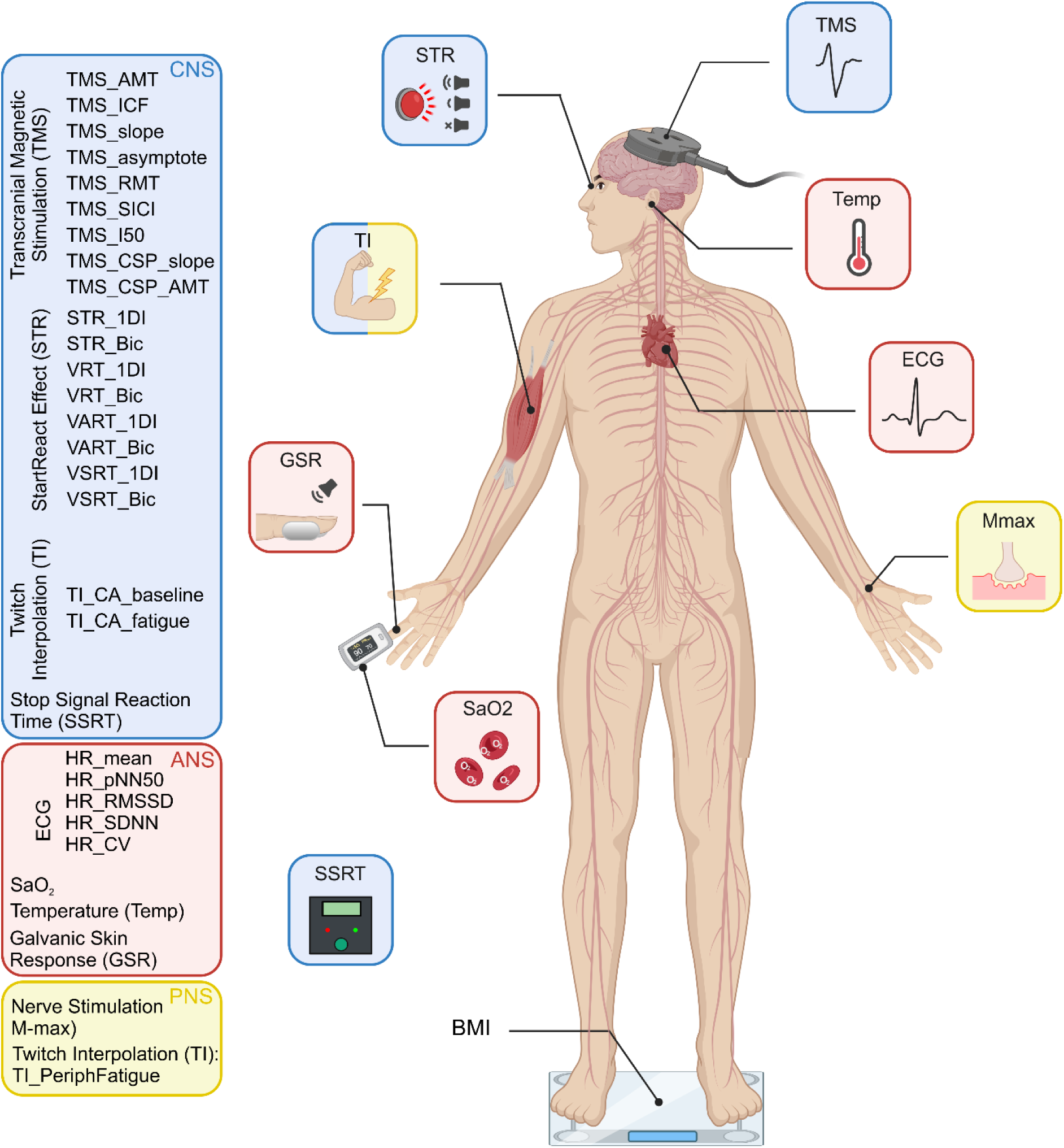
Schematic representation of the electrophysiological measures performed, separated according to which components of the nervous system (CNS, blue; PNS, yellow; and ANS, red) they assessed.

Electromyogram (EMG) was recorded through adhesive surface electrodes placed on the skin over the muscle belly. EMG signals were amplified and filtered (band-width 30-2000 Hz; D360 8-Channel Patient Amplifier, Digitimer, Welwyn Garden City, UK) and then digitized with a sampling rate of 5kHz (CED Micro 1401 with Spike2 software, Cambridge Electronic Design, Cambridge, UK) and stored on a computer for off-line analysis. Where a measurement required a constant contraction, visual feedback of rectified and smoothed EMG activity was provided to the subject via a display of coloured bars on a computer screen, calibrated to the subject’s maximum voluntary contraction (MVC). Participants were then asked to maintain a contraction corresponding to 10% of their individual MVC. Transcranial magnetic stimuli were applied using a figure-of-eight coil through a BiStim 200^2^ stimulator (Magstim, Whitland, UK). The magnetic coil was held to induce electrical currents that flow perpendicular to the presumed line of the central sulcus in a posterior-anterior direction, with the handle pointing backwards and 45° away from the midline. The hotspot was defined as the region where the largest motor-evoked potential (MEP) in the target muscle could be evoked. To ensure a stable coil position during experiments and across sessions, the site of stimulation was marked using a Brainsight neuronavigation system (Rogue Research Inc., Montréal, Canada), which allows online navigation. A Polaris Vicra camera (Northern Digital Inc., Canada) was used to track the coil relative to the head.

#### Peripheral nerve stimulation

Stimuli to peripheral nerves (0.2 ms pulse width) were delivered using a Digitimer DS7AH isolated, constant current stimulator. The size of the maximal M wave was measured by stimulating the median nerve at the wrist and recording EMG from the abductor pollicis brevis (APB) muscle. Stimulus intensity was set to produce a supra-maximal M wave. Ten stimuli at this intensity were delivered and the highest amplitude M wave was used for subsequent normalisation of TMS recruitment curves (see below).

#### TMS recruitment curve

As a measure of motor cortical excitability, the increase in APB MEP with increasing stimulus intensity was used^8^. The active motor threshold (AMT) was defined as the intensity which produced a MEP > 100 µV amplitude in at least 3 out of 6 stimuli, while the participant maintained an active contraction of 10% MVC. The intensity of the stimulation was expressed as a percentage of the maximal stimulator output (MSO). Recruitment curves of increasing intensities in 10% MSO steps were obtained in blocks of ten stimuli per step starting at AMT intensity, until 100% MSO was reached.

Offline analysis measured the size of MEPs from single trials and plotted this versus stimulus intensity. A sigmoid curve could then be fitted to the relationship^8^.

#### Paired-pulse TMS

The hotspot was defined as the region where the largest MEP in the first-dorsal interosseous (1DI) muscle could be evoked with the minimum intensity. Resting motor threshold (RMT) was defined as the minimal stimulus intensity needed to produce a MEP > 100 µV amplitude in at least 3 out of 6 stimuli.

For the test stimulus, TMS intensity was adjusted to elicit MEPs of 1mV amplitude at rest, or to 120% RMT, whichever was lower. The conditioning stimulus intensity was set at 80% RMT.

The conditioning stimulus preceded the test stimulus by 3 or 10 ms and the recorded responses were expressed as a percentage of responses to the test stimulus alone, to measure short-interval intracortical inhibition (SICI) and intracortical facilitation (ICF) respectively^8^. Twenty stimuli for each condition were given in a pseudo-random order.

#### Galvanic skin response

The galvanic skin response (GSR) is a measure of the cutaneous resistance or conductivity, which can be quantified by passing electricity through a pair of electrodes. Variation in skin resistance depends on sweat production, which itself is mediated by the sympathetic system^8^; its habituation may be a relevant measure in assessing cognitive states^8^. The GSR was measured by placing two metal plates on the lateral and medial surfaces of the index finger. Five loud sounds (115 dB, C weighting, 500Hz, 50 ms, 8-8.8s inter-stimulus interval, chosen randomly from a uniform distribution) were played through loudspeakers placed underneath the subject chair, with the subject at rest. The ratio of the GSR response amplitude following the last stimulus compared to the first was used as a measure of habituation.

#### StartReact effect

This paradigm measures reaction time from EMG in response to a visual cue (visual reaction time, VRT), a visual plus quiet auditory cue (visual-auditory reaction time, VART), and a visual plus loud auditory cue (visual-startle reaction time, VSRT). The acceleration of reaction time between VART and VSRT is termed the StartReact effect and assesses connections from the reticulospinal system^8^.

A green light-emitting diode (LED) was located ∼1 m in front of the subject. Participants were instructed to flex their elbow and clench their fist as quickly as possible after the LED illuminated. EMG was recorded from both the first dorsal interosseous muscle (1DI) and biceps muscle, and reaction time measured as the time from cue to onset of the EMG burst. Three types of trial were randomly interleaved (20 repeats per condition; inter-trial interval 6-6.8s, interval chosen randomly from a uniform distribution): LED illumination alone (VRT), LED paired with a quiet sound (80dB, 500Hz, 50ms, VART), LED paired with a loud sound (120 dB, 500 Hz, 50ms, VSRT).

StartReact measurements were performed immediately after the GSR habituation test, ensuring that any overt startle reflex had been habituated by the five loud sounds given in that test. The room lights were dimmed for this test.

Data were analysed offline trial-by-trial using a custom MATLAB program which identified the reaction time as the point where the rectified EMG exceeded the mean + 7 SD of the baseline measured 0-200 ms prior to the stimulus. Every trial was also inspected visually, and erroneous activity onset times (caused, for example, by electrical noise artefacts) were manually corrected. Average VRT, VART and VSRT were calculated for each subject and muscle, together with the amplitude of the StartReact effect, equal to VSRT-VART.

#### Stop-signal reaction time

The stop-signal reaction time (SSRT) task measures the ability to inhibit a response after receiving a GO cue. Participants were asked to respond to a GO cue, but to inhibit their responses if a STOP cue appeared. This can indicate the state of premotor cortical-subcortical areas involved in impulse control^8^ and allows for increased levels of inhibition in the motor system to be tested. The hardware used to measure SSRT in this study was a portable device recently developed that uses Bayesian statistical analysis to improve the reliability of the measure^23^. The battery-powered device consisted of a plastic box which contained a microcontroller and a LCD screen, as well as one green LED, one red LED and a low compliance press button. Participants initiated a trial by pressing and holding down the response button. They were instructed to release this button as quickly as possible when the green LED illuminated (GO trial; 75% of trials).

In 25% of trials, the red LED illuminated 5, 65, 135 or 195ms after the green LED (stop trial) and participants were instructed not to release the button in these trials. Trials were presented in three blocks of 64, with a 60 s break in between each block. Each block consisted of 48 GO trials and 16 STOP trials (four for each delay). Using the distribution of reaction times on the GO trials, and the proportion of successfully inhibited responses, the algorithm calculated the SSRT as described in full in our previous work^23^.

#### Electrocardiogram

A single channel of electrocardiogram (ECG) recording was captured, using a differential recording from left and right shoulder (bandpass 0.3-30 Hz) and stored for offline analysis. The time of each QRS complex was extracted. From these times, the mean heart rate and the pNN50 (a measure of heart rate variability and defined as the proportion of successive intervals which differ by >50ms) were computed. ECG was captured while participants were engaged in the SSRT test (see above), to ensure consistent behaviour across recordings.

#### Twitch interpolation

Post-viral, immune-mediated disorders of the peripheral nervous system cause failure of signal propagation in nerve, ineffective signal transmission at the neuromuscular junction or reduced signal transmission in muscle fibres. If the same occurs in pCF, this would require a stronger voluntary drive to activate muscles and could give rise to a perception of effortful movements^8^. These features were tested before and after a maximal voluntary biceps muscle contraction by supramaximal electrical stimulation of the biceps muscle (twitch interpolation (TI)^24^).

Subjects sat with their arm and forearm strapped into a dynamometer to measure torque about the elbow. The forearm was held vertically in supination, the upper arm horizontal and the elbow was flexed at a 90° angle. Velcro straps held the wrist, forearm and upper arm in place. Thin stainless-steel plate electrodes (30 x 15 mm) covered in saline-soaked gauze were used for electrical stimulation of the biceps muscle, by taping one electrode over the muscle belly and one over its distal tendon. The individual supramaximal stimulus level was set by increasing the intensity until the twitch response (recorded by the dynamometer) grew no further.

Upon receiving an auditory cue, subjects performed a 3s long maximal voluntary contraction (MVC) until a STOP tone sounded. Electrical stimulation to the biceps was given during MVC, 2s after the go cue and at rest, 5s after the stop cue. This sequence was repeated twice, with a 60 s rest period between GO cues. After another 60s rest, another GO cue then indicated to the subject to make a sustained MVC of up to 90s. If exerted force fell below 60% of the initial maximal level, contraction was terminated early. During this long MVC, the biceps muscle was stimulated every 10s. A final three stimuli (inter-stimulus interval 5s) were given at rest.

If a subject truly performs a maximal voluntary contraction, a superimposed electrical stimulus should not be capable of generating extra force. The size of any elicited twitch thus measures a central activation deficit. Stimulation of a fatigued muscle at rest after the long contraction produces a smaller twitch than that seen before the sustained MVC, indicating peripheral fatigue.

Central activation before and after fatigue, as well as peripheral fatigue were computed based on elicited twitches according to the calculations in McDonald, et al. ^24^.

#### Biometric Data

Various biometric measurements were collected from participants. These included blood oxygen saturation, tympanic temperature, height, weight and full body composition (TANITA BC-545N Segmental Body Fat Scale).

#### Analysis

For the per-protocol analysis reported here, only participants meeting minimum adherence (1h of taVNS in at least 50% of days) were included.

Data processing and statistical analyses were conducted using MATLAB (TheMathWorks, Inc., Natick, MA, USA), SPSS (IBM SPSS Statistics for Windows, Armonk, NY: IBM Corp) and R (http://www.r-project.org/).

Descriptive statistics are given as mean ± standard deviation. Because each measure has different units and scales, data were normalized as a Z-score to allow easy comparison of differences. Z-scores were calculated by taking the difference in means of a measure between datasets (pCF vs healthy controls or pCF before vs after 8 weeks of taVNS usage) and dividing it by the standard deviation of the normative dataset (healthy controls or pCF before taVNS usage respectively). This is a measure of effect size and similar to Hedge’s g measure.

To compare the pCF cohort with the healthy controls, unpaired t-tests were used. To compare pCF at baseline with pCF after 8 weeks of taVNS usage, paired t-tests were used. The Benjamini-Hochberg procedure was used to correct for multiple comparisons^25^. Raw (uncorrected) *P*-values are given throughout this report, but only stated as significant if they passed correction for multiple comparisons. Effect size was characterized with Cohen’s *d*.

## Results

Given the very low levels of adherence across participants, we report the results of a per-protocol analysis in those who met a minimum threshold of adherence to the interventions. Participants that fell below minimum usage had on average 27.3 consecutive missed days, and 62.5/120 missed days, with a mean usage per day of 44.7 minutes. Participants that passed minimum usage had an average of 2.7 consecutive missed days, 7.3 total missed days and a mean usage of 149.8 minutes per day. Of the 114 participants recruited, 24 dropped out during the study and a further 31 were excluded from analysis for not meeting minimum TENS usage (see Figure 3B). The following analysis is based on the remaining 59 participants (45 females, 49.8 ± 10.2 years). 24 were randomized to active nVNS, 19 received sham stimulation and 16 were allocated to the placebo group. Participants receiving sham or placebo stimulation for the first 8 weeks crossed over to receive open label active nVNS for the remaining 8 weeks. The control cohort consisted of 52 healthy participants, with no symptoms of fatigue (37 females, 46.1 + 9.3 years).

### Baseline Post-COVID Fatigue in Comparison to Healthy Controls

All baseline measures were compared to the control cohort with an unpaired t-test. 28 out of 46 measures were significantly different between the cohorts after adjusting for multiple comparisons. Figure 5A illustrates results for all electrophysiological measures as a spider plot, normalized as Z-scores and ordered so the greatest difference is located at the top of the figure.

**Figure 5:**
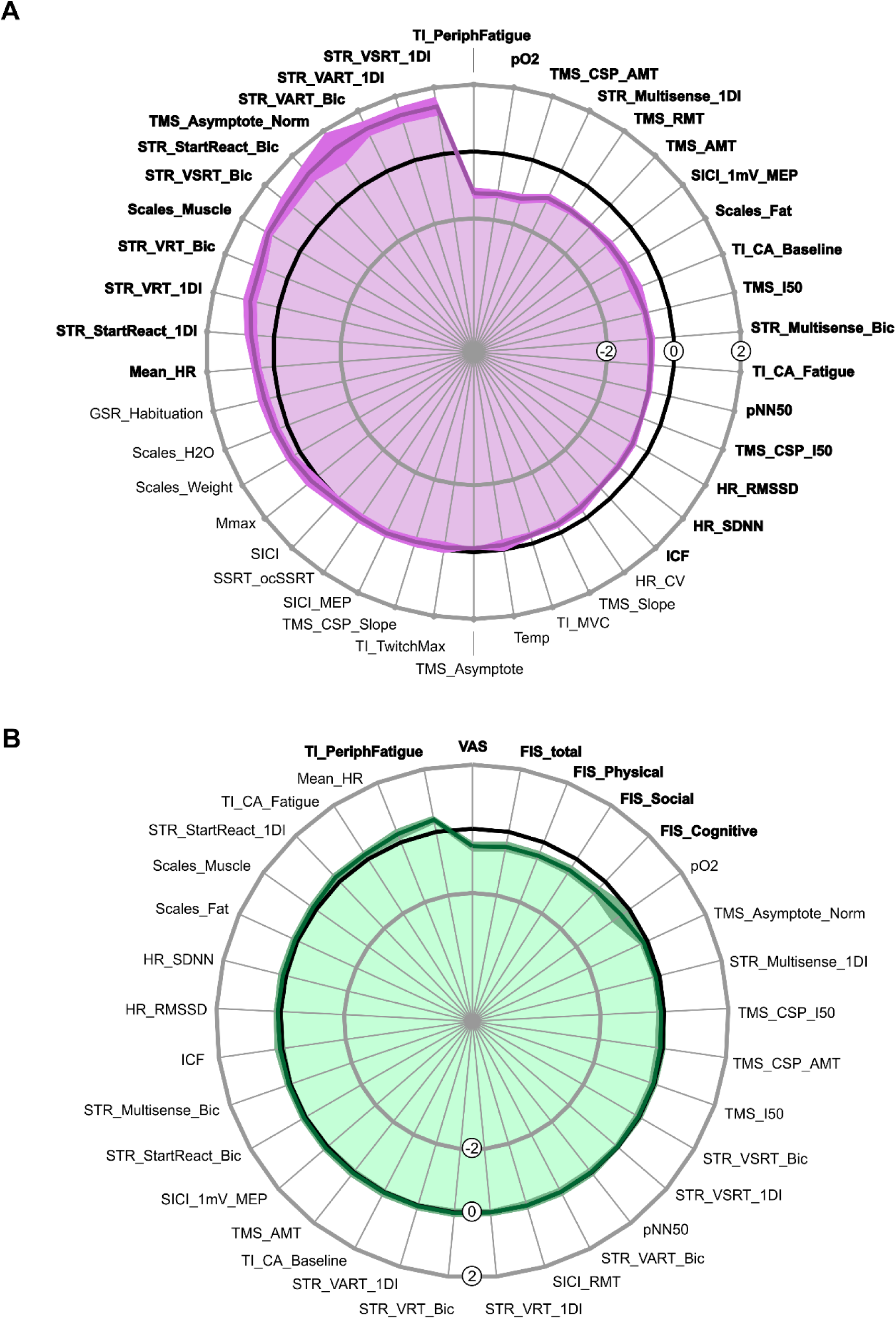
Comparisons of electrophysiological measures**. A:** Differences between pCF and healthy controls. Data for each measure normalized as Z-scores (difference between pCF and control participants, scaled by SD). Black line at zero represents the control cohort, dark purple line the pCF cohort. Purple shading indicates the standard error of the mean difference (calculated by dividing the SD of each measure by the square root of the number of observations available). Measures are ordered so that the greatest difference is located at the top of the figure. Significant differences between the cohorts were assessed using unpaired t-tests. Measures passing the Benjamini-Hochberg correction for multiple comparisons are indicated in bold. **B:** Differences before and after 8 weeks of taVNS. Data for each measure normalized as Z-scores (difference between before and after 8 weeks of using taVNS, scaled by SD). Black line at zero represents the session before using taVNS (session 1 for active nVNS participants; session 2 for sham or placebo participants), dark green line the session after 8 weeks of taVNS usage (session 2 for active nVNS participants; session 3 for sham or placebo participants). Green shading indicates the standard error of the mean difference (calculated by dividing the SD of each measure by the square root of the number of observations available). Measures are ordered so that the greatest difference is located at the top of the figure. Significant differences between the sessions were assessed using paired t-tests. Measures passing the Benjamini-Hochberg correction for multiple comparisons are indicated in bold.

Multiple TMS measures related to cortical excitability were significantly different between controls and pCF (resting motor threshold *TMS_RMT* p<0.001, active motor threshold *TMS_AMT* p<0.001, the intensity yielding 50% of the asymptote response amplitude *TMS_I50* p<0.001, the asymptote of the recruitment curve normalized to M-max *TMS_Asymptote_Norm* p=0.029, active motor threshold of the cortical silent period (CSP) *TMS_CSP_AMT* p<0.001, I50 of the CSP *TMS_CSP_I50* p<0.001), while other TMS measures (the recruitment curve slope *TMS_Slope* p=0.153, the asymptote of the recruitment curve *TMS_Asymptote* p=0.436 and recruitment curve slope of the CSP *TMS_CSP_Slope* p=0.798) showed no difference between the cohorts. Additionally, intracortical facilitation was significantly decreased in pCF (*ICF* p=0.003), but short-interval intracortical inhibition did not show a difference between pCF and controls (*SICI* p=0.746).

We found longer reaction times in pCF versus controls for all measures relating to the StartReact paradigm in both muscles (visual reaction time *STR_VRT_Bic* p<0.001 and *STR_VRT_1DI* p<0.001, visual-auditory reaction time *STR_VART_Bic* p<0.001 and *STR_VART_1DI* p<0.001, visual-startle reaction time *STR_VSRT_Bic* p<0.001 and *STR_VSRT_1DI* p<0.001). The StartReact effect, which measures the acceleration of a visual reaction time by a loud (startling) sound and has been proposed to assess reticulospinal pathways^26^, was also significantly increased in both muscles (*STR_StartReact_Bic* p<0.001 and *STR_StartReact_1DI* p=0.005), while the difference between visual-auditory reaction time and visual-startle reaction time was reduced in pCF versus controls (*STR_Multisense_1DI* p<0.001 and *STR_Multisense_Bic* p<0.001). The stop-signal-reaction time (*SSRT_ocSSRT* p=0.886) was similar for both cohorts.

We also found evidence of autonomic dysregulation in the pCF group, showing lower peripheral blood oxygen saturation (*pO2* p<0.001), increased resting heart rate (*Mean_HR* p=0.010) and lower heart rate variability (*pNN50* p<0.001, *HR_RMSSD* p=0.004, *HR_SDNN* p=0.005). Other measures of autonomic function (tympanic temperature *Temp* p=0.550, galvanic skin response habituation *GSR_Habituation* p=0.073 and variability in heart rate coefficient of variation *HR_CV* p=0.075) did not differ between cohorts.

Maximal voluntary contraction (*TI_MVC* p=0.129) was not significantly reduced in pCF, suggesting no deficit in force production for brief contractions. There also was no difference in their maximal M-wave (*Mmax* p=0.472), or their maximal stimulus-evoked muscle twitch (*TI_TwitchMax* p=0.700). However, after a prolonged maximal contraction, pCF participants had an increased level of peripheral fatigue (size of maximum twitch evoked by direct electrical stimulation of the muscle after a sustained contraction compared with baseline; *TI_PeriphFatigue* p<0.001).

Central activation, which assesses the ability of the CNS to activate muscle maximally voluntarily, was significantly reduced in pCF, either assessed at baseline (*TI_CA_baseline* p=0.002) or after a fatiguing contraction (*TI_CA_fatigue* p<0.001).

In terms of body composition, pCF participants had significantly more muscle mass (*Scales_Muscle* p<0.001) and less fat percentage (*Scales_Fat* p<0.001), whereas there was no difference in water percentage (*Scales_H2O* p=0.091) or actual weight (*Scales_Weight* p=0.119) between the cohorts.

Our results are largely consistent with our previously reported study comparing the same control cohort to a different pCF cohort^8^. However, for the present study we found many more metrics to be significantly different between pCF and controls, possibly because the present pCF cohort had been suffering from the condition for significantly longer than our previous cohort.

### Changes After 8 Weeks of taVNS

The 28 electrophysiological measures that were significantly different in pCF versus control participants at baseline (Figure 5A), as well as the FIS and VAS scores, were then analysed before and after 8 weeks of daily taVNS usage. For this, all pCF participants that met minimum usage were analysed and participants were pooled across the three treatment groups. For participants randomized to active nVNS, the session before taVNS corresponded to session 1 at week 0 and the session after 8 weeks of taVNS usage to session 2 at week 8. For participants randomized to sham or placebo stimulation, the session before taVNS corresponded to session 2 at week 8 (having received only sham or placebo stimulation for the first 8 weeks) and the session after 8 weeks of taVNS usage to session 3 at week 16.

Figure 5B represents results for all measures as a spider plot, normalized as Z-scores and ordered so the greatest difference is located at the top of the figure. Of the 33 total measures (28 electrophysiological measures and 5 questionnaire scores), 6 measures were significantly improved after 8 weeks of taVNS.

Participants improved significantly in all questionnaire scores, including the primary outcome measure of the study (visual analogue scale *VAS* p<0.001), the total score of the fatigue impact scale (FIS_Total p<0.001), as well as all subscores of the fatigue impact scale (FIS_Physical p<0.001, FIS_Social p<0.001, FIS_Cognitive p<0.001).

Participants also had a significantly decreased level of peripheral fatigue (TI_PeriphFatigue p=0.001), which compared to subjective questionnaires provides an objective electrophysiological measure of improvement.

All other measures remained unchanged after 8 weeks of taVNS usage.

### Differences in treatment groups

Figure 6 compares these 6 significant measures between sessions and treatment groups.

**Figure 6:**
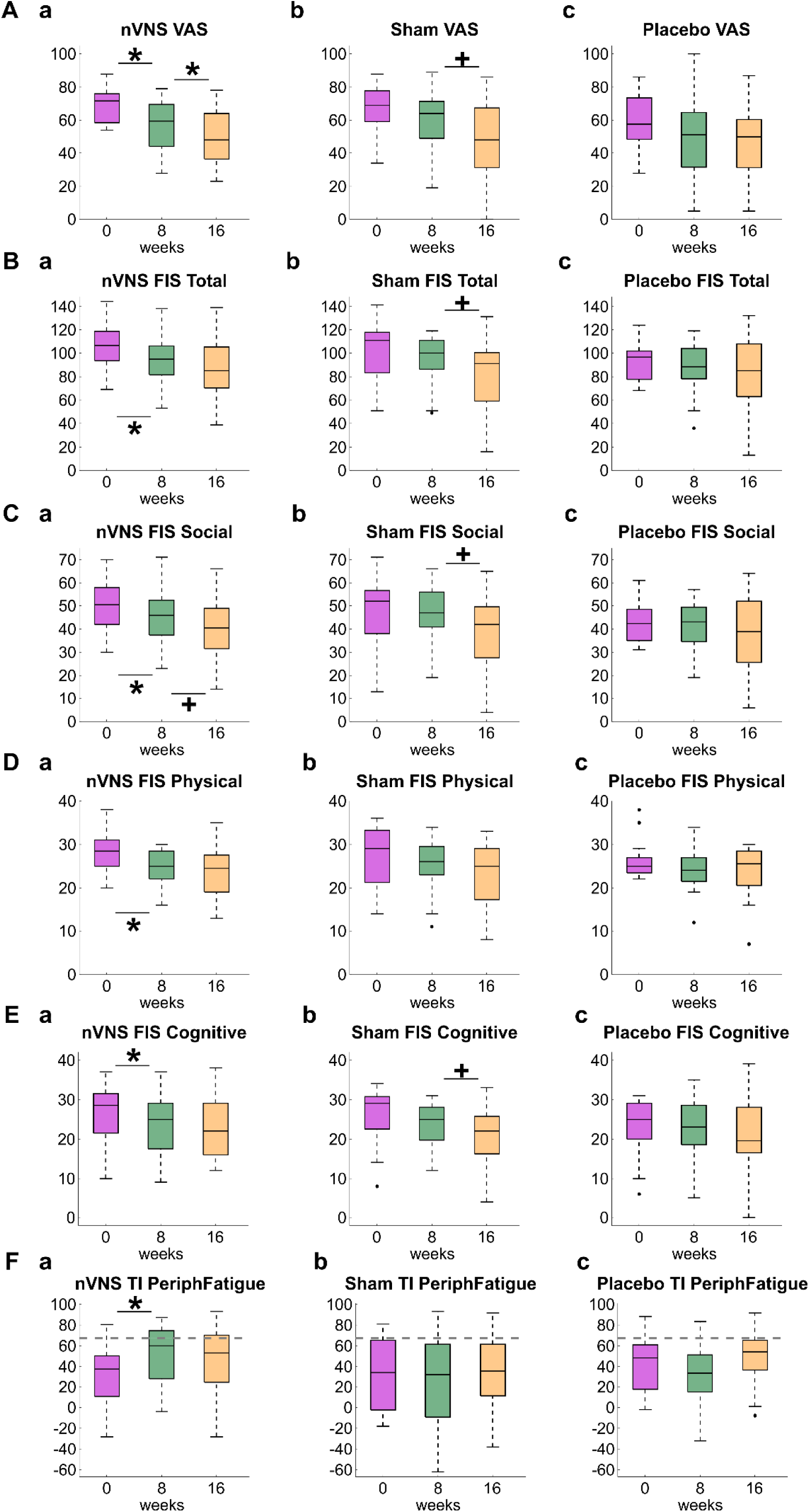
Box and Whisker plots for the measures that showed significant changes after 8 weeks of taVNS usage for the active nVNS (left column), sham (middle column) and placebo (right column) group. Measures were compared between week 0 (purple), week 8 (green) and week 16 (mustard) with paired t-tests. Asterisks indicate significant changes that pass correction for multiple comparisons. Crosses indicate trends (p<0.05) that did not pass multiple comparisons. Note that all significant changes are in the active nVNS group.

A repeated measures ANOVA showed no significant interaction between group and session for the visual analogue scale (*VAS* F(4,110)=1.516, p=0.203), FIS scores (*FIS_Total* F(4,112)=0.727, p=0.575, *FIS_Social* F(4,112)=0.770, p=0.548, *FIS_Physical* F(4,112)=0.680, p=0.607, *FIS_Cognitive* F(4,112)=0.691, p=0.600), or peripheral fatigue (*TI_PeriphFatigue* F(4,104)=2.884, p=0.026; not significant after correcting for multiple comparisons).

Between week 0 and week 8, only the active nVNS group showed significant improvement in VAS (11.9 ± 17.8 points reduction, p=0.003, d=0.939; Figure 6Aa). Although the sham and placebo group display the same direction of change (8.0 ± 13.4 points reduction; Figure 6Ab & 8.0 ± 21.0 points reduction; Figure 6Ac), this change did not reach significance in either of the control groups (sham VAS p=0.018, d=0.513; not significant after correcting for multiple comparisons, placebo VAS p=0.141, d=0.384).

Active nVNS for 8 weeks also significantly improved all FIS scores (*FIS_Total* 11.1 ± 17.5 points reduction, p=0.005, d=0.544, Fig. 6Ba; *FIS_Social* p=0.026, d=0.426, Fig. 6Ca; *FIS_Physical* p<0.001, d=0.849, Fig. 6Da; *FIS_Cognitive* p=0.020, d=0.339, Fig. 6Ea). This improvement was not seen in the sham (*FIS_Total* 5.2 ± 12.1 points reduction, p=0.077, Fig. 6Bb; *FIS_Social* p=0.421, Fig. 6Cb; *FIS_Physical* p=0.023, Fig. 6Db; not significant after correcting for multiple comparisons, *FIS_Cognitive* p=0.072, Fig. 6Eb) or placebo (*FIS_Total* 4.6 ± 19.9 points reduction, p=0.367, Fig. 6Bc; *FIS_Social* p=0.427, Fig. 6Cc; *FIS_Physical* p=0.104, Fig. 6Dc; *FIS_Cognitive* p=0.758, Fig. 6Ec) group.

The active nVNS also showed a significant decrease in peripheral fatigue (*TI_PeriphFatigue* p=0.004, d=-0.531, Fig. 6Fa), which was unchanged after 8 weeks of sham (p=0.876, Fig. 6Fb) or placebo (p=0.171, Fig. 6Fc) stimulation.

Upon crossing over to receive active stimulation, the sham group showed a trend of improvement in VAS (p=0.009, Fig. 6Ab), total FIS score (p=0.044, Fig. 6Bb) and social FIS score (p=0.030, Fig. 6Cb); however, this was not significant after correcting for multiple comparison. The placebo group did not show any significant changes in fatigue PROs after crossing over to receive active nVNS (between week 8 and week 16). This is presumably because, after adjusting for dropouts and participants not meeting minimum usage, the placebo group was left with fewer participants (n=16) than the active nVNS (n=24) or the sham (n=19) groups.

There was additional improvement in VAS when using active taVNS for another 8 weeks (16 weeks total), though it was less pronounced than over the first 8 weeks (decrease of 7.0 ± 10.4 points, p=0.003, d=0.437; Fig. 6Aa). There was no further reduction in FIS scores or peripheral fatigue after 16 weeks of taVNS (Fig.6Ba & Fig. 6Fa).

## Discussion

This study provides evidence that taVNS may offer a non-invasive and non-pharmacological approach to improve pCF, even in patients who have had ongoing symptoms for years. Significant improvements were only seen in the active nVNS group and not in the sham or placebo group. However, adherence posed a considerable problem, and these significant changes were only seen after excluding participants not meeting a minimum usage criterion.

Apart from assessing the effects of taVNS on pCF, we were also able to add to previous observations on physiological differences between people suffering post-COVID fatigue and healthy controls.

Over half of the measures tested (28 out of 46) were significantly different between pCF and healthy controls. These included changes in cortical excitability, autonomic regulation, muscle physiology and even body composition. Fatigue is a multifaceted phenomenon with cognitive, neuromuscular, autonomic, psychological and social factors.

Cognitive fatigue is a decline in cognitive functions, including attention, working memory, judgment and memory recall^27^. We found reaction times were significantly slower in pCF, indicating diminished cognitive processing^28^. On the other hand, the StartReact effect, which measures the acceleration of a visual reaction time by a loud (startling) to assess reticulospinal pathways, was increased in pCF. This may be consistent with homeostatic compensation for increasing peripheral fatigue and weakness.

Neuromuscular fatigue refers to reduced muscle force generation; it has both peripheral and central contributions. Peripheral fatigue is characterized by a failure of the peripheral nervous system whereas central fatigue reflects a decline in the CNS ability to activate muscles maximally.

Several changes were found centrally. Cortical excitability was increased in the pCF cohort, with lower motor thresholds to TMS and larger MEP sizes. The CSP was also reduced, similar to another study which showed reduced CSP after a fatiguing motor task in post-COVID patients^29^. These findings point to cortical over-excitability, but conversely, intra-cortical facilitation (ICF), a measure of intracortical glutamatergic function, was reduced in pCF. Short-intra-cortical inhibition (SICI), which measures GABA_A_ function, was not different from healthy controls. This replicates findings from our previous study^8^. Collectively these cortical changes might underlie that impaired central activation in pCF.

People with pCF showed no difference in maximal voluntary contraction or stimulus-evoked muscle twitch at rest. Myopathic changes only became apparent after a sustained contraction, when the ability of muscle to produce force in response to electrical stimulation was significantly reduced in pCF subjects. In fact, this measure of peripheral fatigue showed the greatest difference between cohorts of any measure.

Acute SARS-CoV-2 infection can impair skeletal muscle function in multiple ways. Viral entry into host cells via the ACE-2 receptor and TMPRSS2 protein can cause direct muscle cell damage^30^. SARS-CoV-2 may impair mitochondrial function in muscles directly^31^, even in individuals suffering from PCS that have not been hospitalised^5,6^. Additionally, infection promotes a pro-inflammatory muscle environment^32,33^. Under physiological conditions, exercise-induced myokines exert anti-inflammatory effects; however, during SARS-CoV-2 infection, altered myokine signalling sustains inflammation^32,33^, which in turn impairs mitochondrial function^34^.

pCF subjects had a greater muscle mass per body weight than healthy controls. We speculate that this could be caused by muscle metabolic changes. Fast-twitch (FT) muscle fibers are typically larger and denser than slow-twitch (ST) fibers, contributing to greater muscle size and power^35^. Over time, to counteract peripheral fatigue, pCF sufferers could increase the fraction of FT fibers, which have lower mitochondrial density than ST fibers^36^. This shift could increase overall muscle mass and yet the increased reliance on FT fibers would lead to greater fatigue. This speculative suggestion needs to be verified by measuring muscle fiber composition directly in a future study.

An increased body mass index (BMI) is sometimes listed as a risk factor for developing PCS^37^. However, most studies on PCS have focussed on hospitalized patients; information on non-hospitalized patients is scarce. While higher BMI is clearly correlated with disease severity and therefore hospitalization^38^, there is no strong proof that it predisposes people with milder infections to PCS. Indeed, we did not observe a difference in weight between cohorts. pCF subjects had a smaller body fat percentage than healthy controls; it is impossible to say if that represents body composition pre-COVID or a change resulting from the condition.

Autonomic dysregulation has long been associated with chronic fatigue^39^ and multiple studies have reported autonomic dysfunction after COVID-19^40,41^. Supporting this, we found multiple abnormalities in autonomic function. People with pCF had significantly increased resting heart rate and lower heart rate variability, reflecting reduced parasympathetic activity and/or sympathetic overdrive^42^. We also found lower peripheral blood oxygen saturation, although this might be the result of persistent pulmonary injury or vasculopathy, which is often found after SARS-CoV-2 infection^43,44^. Other measures of autonomic function, such as tympanic temperature and galvanic skin response habituation were unchanged for this cohort, contrasting with increased temperature and decreased GSR habituation found in our previous study^8^.

Autonomic dysregulation is indicative of an overall reduction in vagal activity and a shift towards sympathetic overactivation. Vagus nerve stimulation could therefore plausibly help to restore autonomic balance. The vagus nerve consists of approximately 80% afferent and 20% efferent axons^45,46^. Stimulation of the auricular branch primarily engages afferent pathways, which project to the nucleus of the solitary tract, triggering a cascade of neurophysiological responses that may potentially produce therapeutic effects^47,48^. taVNS activates largely the same neural pathways as invasive VNS^49^. Proposed mechanisms for therapeutic benefits include heart rate and blood pressure control^50,51^, prevention of inflammation via cholinergic anti-inflammatory pathways^52,53^, dampening the body’s stress response^54^ and restoration of autonomic function^55^.

Recently, non-invasive VNS has successfully been used in acute COVID-19 patients to reduce biomarkers of inflammation^18,19^. In a pilot study using taVNS for long COVID symptoms, 20 people suffering from PCS reported a decrease in symptom severity, including fatigue, after 10 days of daily taVNS^21^. Another pilot study used 10 days of daily taVNS in 24 female long-COVID patients, and found significant improvements in various cognitive functions, anxiety, depression and sleep^20^. However, both pilot studies did not include a control group for comparison. Although one study using at home taVNS in PCS patients did include a randomized sham control^56^, with only 6 patients per treatment arm it was underpowered.

The present work builds upon these previous findings. After 8 weeks of at-home taVNS, participants that met the minimum usage showed a significant reduction in fatigue measured on a VAS. This change was only significant in the active nVNS, and not sham or placebo group. No consensus currently exists on how to determine minimal clinically important difference (MCID). Previous approaches include the SD/2 rule^57^, 0.2 SD^58^, 0.3 SD^59^, or using 5-10% of the instrument range^60^. By any of these methods, the reduction of fatigue we observed is clinically significant, as corroborated by the large effect size. There was additional improvement when using active taVNS for another 8 weeks (16 weeks total), though it was less pronounced than over the first 8 weeks (decrease of 7.0 ± 10.4 points). Using all approaches except the 10% instrument range method, this would still count as a clinically significant change, but the effect size was small.

After switching to active nVNS at week 8, the placebo and sham groups saw some improvement in the VAS. While this change had a medium effect size, it was not significant after correction for multiple comparisons.

The active nVNS group also improved across all domains of the Fatigue Impact Scale. Again, no significant change occurred in the sham or the placebo groups. There was no further reduction in FIS scores after another 8 weeks of taVNS, indicating the effect might have reached a plateau. Similar to the VAS results, the sham group showed a trend in improvement of total FIS score after switching to active nVNS at week 8.

In addition to subjective perception of fatigue, 8 weeks of active taVNS also significantly improved peripheral muscle fatigue.

We would expect to replicate the changes seen in the active group in the sham and placebo group, once the participants switched to the active intervention in weeks 8-16. The sham group did indeed show promising trends in several measures, including improvements in VAS and FIS scores, but the placebo group did not. The discrepancy in the three groups may be due to small participant numbers.

It is notable that the main measures showing improvement after 8 weeks of taVNS (FIS score, peripheral fatigue) were the same as those which recovered spontaneously in many participants after 12 months in our previous study^22^. It is unclear if the spontaneous improvements reflect a recovery back to healthy baseline function or a process of adaptation. In line with the Bradford-Hill criteria for causality^61^, the observation that targeted perturbation of the system leads to measurable, correlated physiological and symptomatic changes strengthens the argument that vagal dysfunction is not merely correlated with but actively contributes to pCF. Post-COVID fatigue might therefore more accurately be termed ‘Sick Vagus Syndrome’.

The fact that peripheral muscle fatigue measured by electrical stimulation was the only physiological measure to demonstrate significant changes after taVNS does not necessarily mean other measures were not affected. Rather, this stimulation-based measure may have especially low inter-measurement variability, potentially making it a highly sensitive, quantifiable and relatively low-noise biomarker of fatigue. Clinically, peripheral fatigue could be a valuable early marker of recovery or treatment response, with broader changes in cortical and autonomic domains possibly emerging over longer follow-up or in larger samples.

Adherence was the biggest limitation in this study, with a third of the participants not meeting the minimum usage criterion. Although low, this matches the rate of adherence to medication therapies at around 50% in high-income countries^62^. Additionally, adherence is known to be higher among patients with acute compared with chronic conditions^63–65^. Little is known about adherence for at-home stimulation therapies. None of the published studies applying at-home taVNS interventions reported adherence, presumably because the devices used to deliver the stimulation had no means of reporting usage. Here we incorporated a custom-made data logger which collected data on device usage. Adherence is a vital factor to evaluate the efficacy of any therapy, since participants not using the device will disproportionally dilute overall group effects of the intervention. Conflicting reports on the efficacy of taVNS as a treatment in chronic conditions might well be the result of divergent adherence levels across the different studies, leaving the interpretation of these typically heterogeneous populations inconclusive. Notably, our predefined threshold for acceptable device use – at least one hour on only 50% of days – was deliberately lenient. Observing significant effects even at this minimal level suggests that more consistent or prolonged stimulation may yield substantially greater therapeutic benefit, highlighting the need to investigate optimal dosing.

In this study, we observed significant improvements after 8 weeks of daily taVNS that was not seen after sham or placebo stimulation. The improvements notably include the primary outcome of fatigue VAS, perceived impact of fatigue in all FIS scores, as well as the objective physiological measure of peripheral fatigue. taVNS may offer a promising non-invasive and non-pharmacological approach to manage pCF. Future investigations should focus on potential long-term retention of benefits and address the adherence problem by finding reasons why participants struggled to keep up with the treatment. Subsequent research is also needed to understand the biological mechanisms of taVNS, which would allow better tailoring of therapeutic approaches to pCF.

## Supporting information

Supplementary information: Data logger design

## Data Availability

All data produced in the present study are available upon reasonable request to the authors

