## Supplementary information: Data logger design for "Percutaneous Auricular Nerve Stimulation for Treating Post-COVID Fatigue (PAuSing-pCF)"

The MultiSTIM TENS device is a CE-marked Medical Device; to maintain this certification, no modification was made to the device itself. Sensing whether the device was turned on and stimulating was achieved by gently separating the two wires of the cable leading from the stimulator, and wrapping them each twice around a ferrite rod (8 mm diameter, 21 mm length). The two wires were wrapped in opposite directions (one clockwise, one anti-clockwise), so that opposite directions of current flow would produce the same direction of magnetic flux. Around the same rod were wrapped 30 turns of enamelled copper wire. Whenever the stimulator delivered a stimulus pulse, the rapid change in current at the beginning and end of each pulse induced a brief voltage spike in the 30-turn coil.

A schematic of the monitor circuit is shown in Supplementary Figure 1. This was constructed on a double-sided PCB, size 48x12 mm, with IC2 and the connector for BAT1 on the underside. The ends of the 30-turn coil described above were soldered onto the PCB forming L1. Voltage spikes from the coil are fed to the comparator IC2, where they are compared to a fixed voltage threshold of around 25 mV generated by the potential divider formed by R1 and R2. Negative spikes from L1 are shorted via D1 to protect the input of IC2. The output of IC2 comprises very brief (~10 ns) pulses. Detecting these without further conditioning would require a high sampling rate, which would incur high power consumption. To avoid this, the comparator output is fed to the trigger input of retriggerable monostable multivibrator IC3, which is set via C1 and R3 to have an output pulse width of approximately 46 ms. This has the effect of widening the detection pulses. At any stimulation frequency above ~21 Hz, the output of IC3 will be continually high, because it will be retriggered by the next stimulus pulse before the output goes low.

The system is controlled by a ATTINY85 microcontroller (IC1). On restarting, terminal PB3 is configured to be an output and set low, thereby powering down the detection circuit involving IC2, IC3 and the potential divider R1/R2 and greatly reducing power consumption. IC1 then enters low power mode; it wakes every 8.56 s to increment a counter. If the counter has not yet reached 70, the device immediately returns to low power mode. Once a count of 70 is reached (cycle time close to 10 minutes), output PB3 is set high, powering up the detection circuit. After a delay of 1 s to allow for startup transients, the output of IC3 is read via terminal PB4. If it is high, an internal counter is incremented. The ATTINY85 then returns to sleep mode. After 24 such cycles, lasting 4 hours, the count is written to internal EEPROM memory, and the count reset. The device thus provides a count between 0 and 24 of how many times over a 4 hour cycle stimulation has been detected. The ATTINY85 has 512 bytes of EEPROM storage; only one byte is needed for each count, allowing the system to run for over 11 weeks.

The system is powered by a CR1220 coin cell battery with 38 mAh rated capacity. During the sleep state, power consumption is 5 µA. When the device makes a measurement every 10 minutes, it wakes for 1.008 s, with a current consumption of 3.76 mA. This gives an average current of 11.3 µA, and suggests a potential run time of 20 weeks within the rated battery capacity. In practice the longest possible run time is therefore limited by the available internal storage, and not by the battery.

Connector CON1 serves two purposes. It is wired to be compatible with the In Service Programming (ISP) standard used by Arduino boards; it is used to download the program to the ATTINY85 when the PCB is first made. Programming also wipes the EEPROM storage, allowing a device to be reused for another subject. Secondly, CON1 allows data download. A cable links PB0 of IC1 via CON1 to the data received terminal of a USB-serial converter module (TTL-232R-3V3-WE, Future Technology Devices International Ltd, Glasgow, UK). The same cable also links PB1 of IC1 via CON1 to ground. The cable is connected to the PCB with the power turned off via switch SW1; it is then powered on. The microcontroller program detects the startup ground state of PB1, and initiates a serial data transfer at 9600 baud of all counts stored in the EEPROM storage.

The PCB was placed in a small box situated within the handle of the MultiStim TENS device, as shown in Fig. 3Aa. The device was prepared for issue to a participant by wiping it using the programmer, and it was turned off using slide switch SW1. Just before issue, it was turned on, and the time noted. This allowed the four-hour count data to be aligned to calendar days as in Fig. 3C.

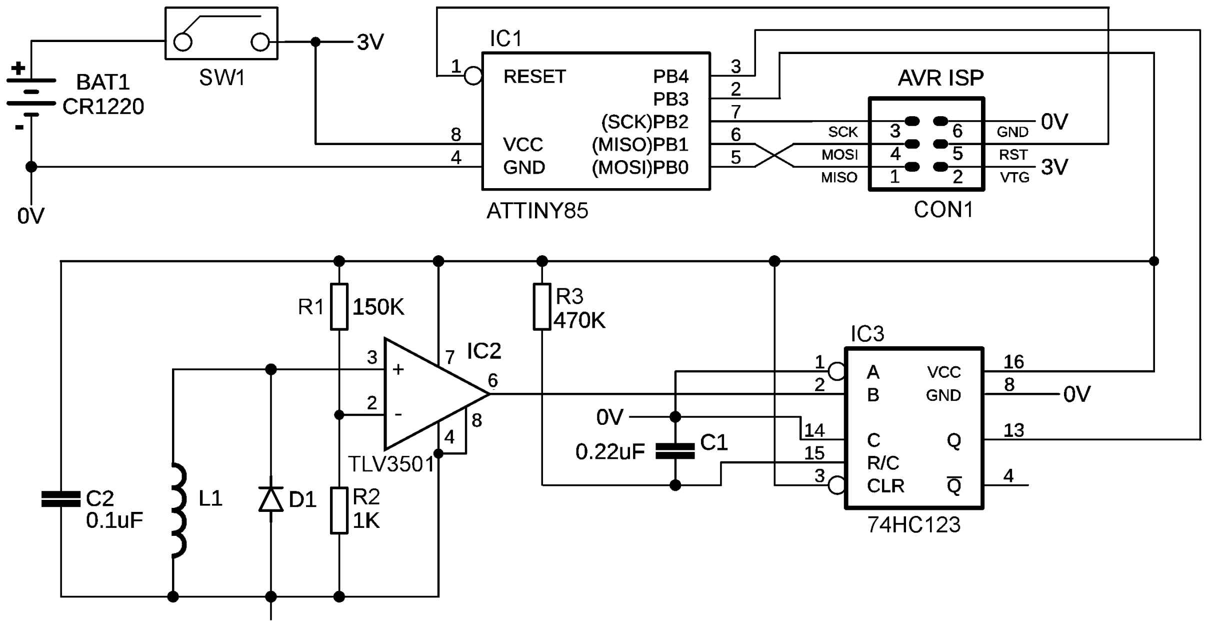


**Supplementary Figure 1.** Schematic circuit diagram of the usage monitor.
